# Projection of COVID-19 Pandemic in Uganda

**DOI:** 10.1101/2020.04.02.20051086

**Authors:** Fulgensia Kamugisha Mbabazi

## Abstract

COVID–19 (Corona Virus) is caused by Severe Acute Respiratory Syndrome Corona Virus 2 (SARS–COV–2). The virus that was first discovered in China Wuhan Province about 3 months ago (first cases were reported in Wuhan on December 31st 2019) has spread world wide. The six (6) top countries (excluding China) most affected so far include; USA, Italy, Spain, Germany, France and Iran. With Italy showing the highest death toll.

In Uganda where it was discovered on 19/3/2020 with one (01) case has just in nine (9) days, increased to thirty (30) infected individuals. This model is a wake up call over the rate at which COVID–19 is likely to spread throughout the country. Thus it is a guide for policymakers and planners to benchmark on for solutions to this deadly virus.

## 1 Introduction

COVID–19 is caused by Severe Acute Respiratory Syndrome Corona Virus 2 (SARS–COV–2) [1]. Corona viruses are widely distributed in many different species of animals, including bats, cattle, cats, birds, and camels [4]. The type of pneumonia caused by the 2019 novel corona virus disease (COVID–19) is a highly infectious disease, and the ongoing outbreak was declared by World Health Organization (WHO) as a global public health emergency on 1/2/2020 [2].

An individual may fail to differentiate between symptoms caused by COVID–19 and a common cold or flu because symptoms are alike. COVID–19 symptoms include: fever and cough that may advance to a severe pneumonia triggering shortness of breath [13]. From the time of exposure/contact to virus, symptoms may take up to two (2) weeks [1-14 days] to show up [11]. In severe cases the disease may consequently lead to death [12].

The top 6 countries reporting the most outside China, globally, Table 1 and Table 2 [9]. The mortality due to COVID–19 varies from country to country, the age and health of the population seem to be aged above 60 years old [6]. Confirmed COVID–19 stand at 657,434 and 30,420 (CFR 4.6%) related deaths have been reported [9]. As of 28 March 2020, 46 African countries have contracted the disease, 8 countries are virus free and there are 3,977 cases, 117 (CFR 3%) deaths, and 286 recoveries across the continent [10].

**Table 1:**
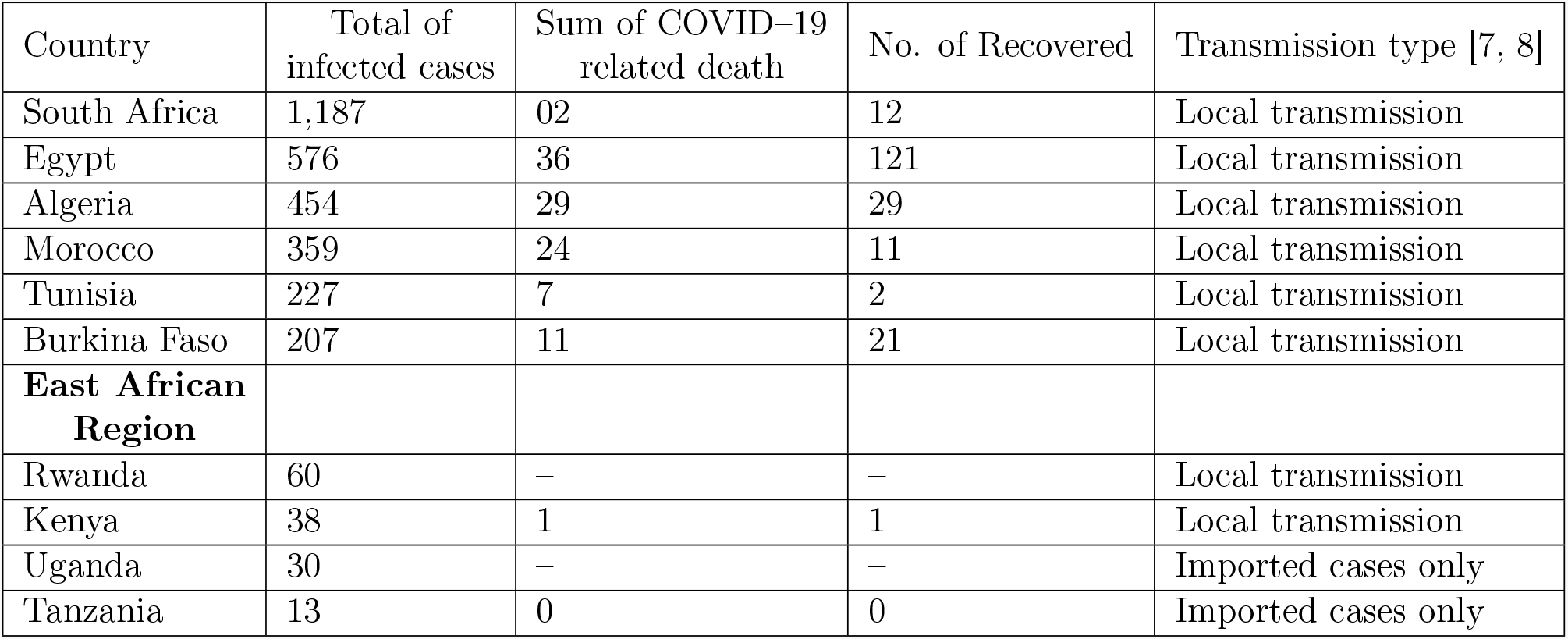
Confirmed COVID–19 Cases Reported in Africa as per 28^*th*^ March 2020, [9]

**Table 2:**
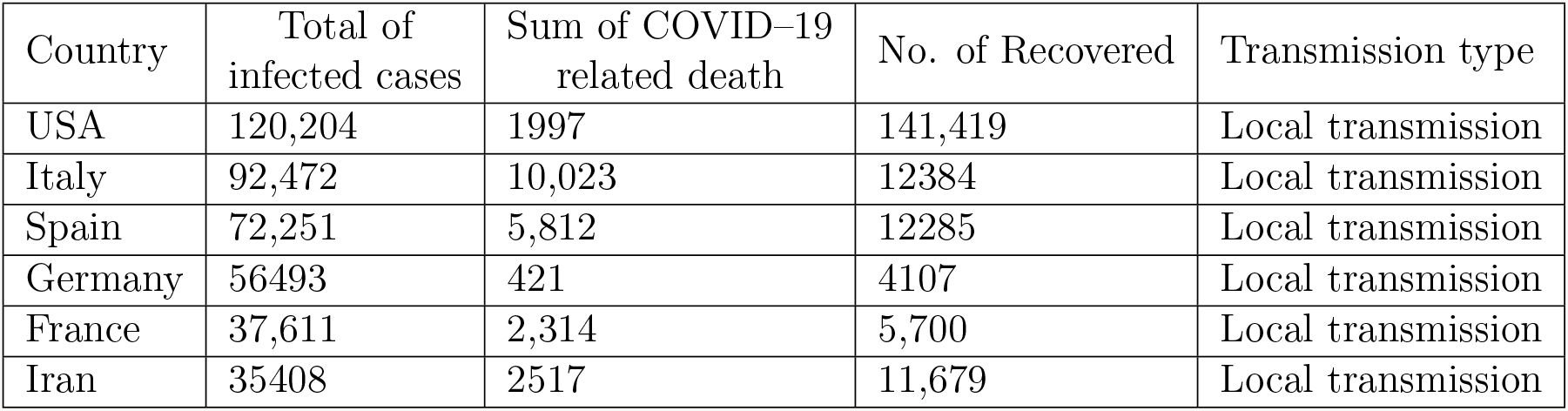
Top six (6) confirmed COVID–19 Cases Reported outside Africa as per 28^*th*^ March 2020, [9]

In Uganda the trend of infected cases is on an increase Table 3. Immunocompromised individuals are likely to be more affected. It should be noted that as of 31^*st*^ March 2020, no reported COVID–19 related death and no Recovered

**Table 3:** Confirmed COVID–19 Uganda cases as per 31^*st*^ March 2020, as announced by Ministry of Health Uganda

| Date | New cases | Total number of infected cases | Transmission type |
| --- | --- | --- | --- |
| 19/3/2020 | 01 | 01 | Imported cases only |
| 21/3/2020 | 8 | 9 | Imported cases only |
| 24/3/2020 | 5 | 14 | Imported cases only |
| 27/3/2020 | 4 | 18 | Imported cases only |
| 28/3/2020 | 5 | 23 | Imported cases only |
| 29/3/2020 | 10 | 33 | Imported cases only |
| 31/3/2020 | 11 | 44 | Local transmission & Imported cases |

## 2 The exponential growth model for projecting number of infections in Uganda

### Assumptions

i. No COVID–19 related death has occurred in Uganda as per the records
ii. The rural population is unaware of the existing control measures
iii. In two days 9 infected individuals are tested positive and quarantined

### 2.1 The Model

Let the infected population at time *t* be *I*(*t*) = *I*. We assume the rate of increase of the infected population 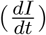 to be directly proportional to the infected population (I).

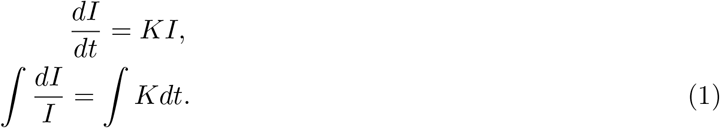

From eqn (1),

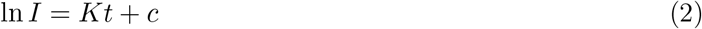

. We note at the beginning of the epidemic Uganda (East Africa–Sub–Saharan Africa) reported 9 infected individuals (2 days) [I(2)=9]. Current information from Ministry of Health–Uganda (25/3/2020) shows an increase by 5 more infected cases, rising to fourteen (14 infected individuals)/ cases in four days(I(4)=14) [5].

From eqn (1) we have

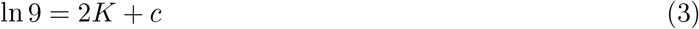

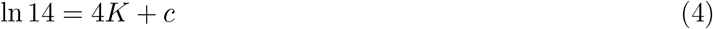

Solving the above equations

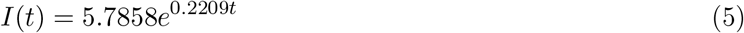

Therefore, from eqn (5) we observe in 2 weeks[14 days]: *I*(14) ≈ 127 infected individuals. In a month (30) days, *I*(30) ≈ 4, 369 infected individuals.

From Figure 1 above, the trend of infected individuals is observed to increase exponentially. In 14 days, the number of infected individuals is expected to be 127 if no control measures are taken.

**Figure 1:**
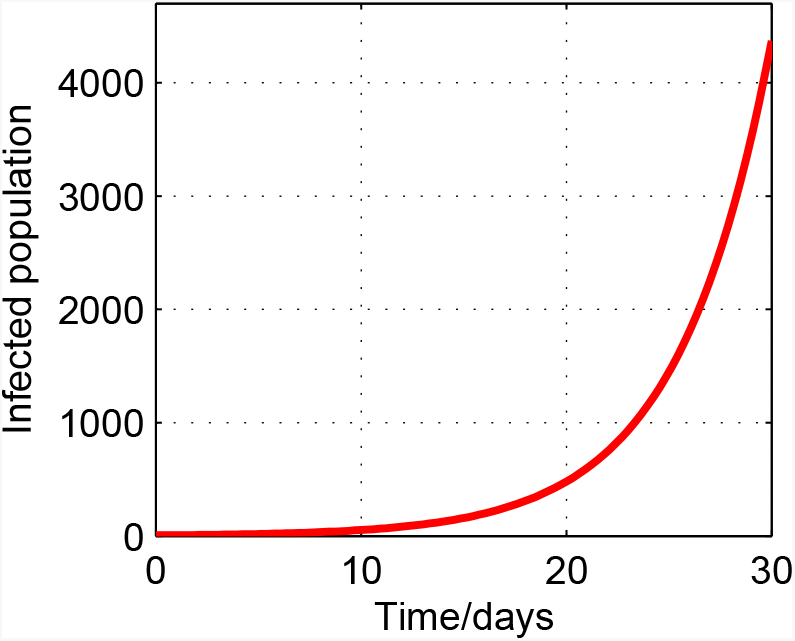
The exponential projection of the number of infected individuals due to COVID–19 in Uganda without interventions (2020)

## Conclusion

With such increasing infected cases, the susceptible population should follow the Ministry of Health guidelines and the advice given by His Excellency Y.K. Museveni the President of the Republic of Uganda. Awareness campaigns [door to door] should be extended to rural populations that have no access to media [Televisions, Radios, Newspapers e.t.c]

No delay in seeking medical care by infected individuals as COVID–19 has a high transmission rate and there is a likelihood of co–infections with other diseases such as Pneumonia (spread by pneumococcus bacteria and influenza A virus, presenting same symptoms), malaria, Tuberculosis and HIV–AIDS.

## Data Availability

The data supporting this exponential model is from the daily updates on COVID- 19, in the world, and they have been suitably cited as references in this paper in Table 1, Table 2 and Table 3. Parameter values have been estimated using data released on a daily basis from the Ministry of Health Uganda.

https://www.africanews.com/2020/03/24/africa-outbreak-brief-10-coronavirus-disease-2019-covid-19-pandemic/

https://www.worldometers.info/coronavirus/

https://www.africanews.com/2020/03/28/coronavirus-africa-46-countries-reporting-a-total-covid-19-3977-cases-117-deaths-and-286-recoveries//

## Acknowledgment

The Author would like to thank Dr. Yahaya Gavamukulya, Busitema University, Uganda and Mr. Kamugisha Joram Kembo for the technical support offered in the preparation of this manuscript.

## Data Availability

The data supporting this exponential model is from the daily updates on COVID–19, in the world, and they have been suitably cited as references in this paper in Table 1, Table 2 and Table 3. Parameter values have been estimated using data released on a daily basis from the Ministry of Health Uganda.

## Conflicts of Interest

The author declares that she has no conflicts of interest.

## Notes

### Competing Interest Statement

The authors have declared no competing interest.

### Funding Statement

This work is so far being undertaken willingly, with no funding from any source

